# Influence of Environmental Exposures on T Follicular Helper Cell Function and Implications on Immunity: a comparison of Bangladeshi and American Children

**DOI:** 10.1101/2024.11.29.24318102

**Authors:** Dana Van Fossen, Hyunjae Cho, Lisa Wagar, Jennie Z. Ma, Rashidul Haque, Mark Davis, William A. Petri

## Abstract

T follicular helper (Tfh) cells are crucial for B cell activation and subsequent antibody production. This functionality is influenced by surface markers such as CD40L, a costimulatory factor which promotes B cell activation, and CD57 which is a well-known marker of senescence. This study examined age-specific differences in Tfh cell function in Bangladeshi and American children. At age two, Bangladeshi children displayed impaired CD40L upregulation and significant CD57 downregulation upon stimulation. These patterns, not observed in American children of the same age, suggested an exhaustion-like phenotype potentially driven by environmental factors. Predictors of Tfh cell response to stimulation were analyzed using Random Forest and Generalized Estimating Equations (GEE) models. Exclusive breastfeeding duration, antibiotic treatments, diarrheal episodes, and malnutrition were identified as variables that significantly impacted the Tfh response to stimuli. To assess Tfh cell ability to promote antibody responses, we correlated Tfh functionality with antibody concentration post-vaccination and in response to infection with Cryptosporidium, an endemic apicomplexan parasite. Increased CD40L expression upon stimulation correlated positively with anti-Poliovirus type 2/3 neutralizing antibody and anti-Cp17 (a Cryptosporidium sporozoite antigen) IgA concentrations. In contrast, increased CD57 expression was significantly correlated with decreased anti-Cp17 IgA. This indicates that an activation-supportive phenotype (CD40L+) may be more effective in promoting immunity than a senescent phenotype (CD57+). Together, these findings suggest that early-life environmental exposures may program Tfh cell functionality, impacting immune response potential in settings with high pathogen exposure.

**IMPORTANCE:** T follicular helper (Tfh) cells are upstream mediators that shape the humoral immune response to specific antigens. The generation of an effective memory response to infection is vital to prevent subsequent reinfections. However, in areas with high burdens of exposure to infections, such as the urban community from Bangladesh studied here, children are consistently exposed to inflammatory pathogens. Specific environmental exposures significantly influenced Tfh cell activation and senescence phenotypes. Additionally, Tfh cell responses correlated with antibody concentrations following vaccination or infection, indicating that environmental factors may play a critical role in shaping effective immunity in early childhood.

## INTRODUCTION

T follicular helper (Tfh) cells, a subset of CD4+ T cells (identified as CXCR5+, PD-1+), play a pivotal role in the activation of B cells, leading to class switching and somatic hypermutation, resulting in high affinity antibody production.^1,2,3^ As such, Tfh cell functionality is a key component of immune competency, particularly in early childhood when the immune system is developing and adapting to first-time environmental exposures and mounting protective responses after vaccination.^4,5^

The functional capacity of Tfh cells is, in part, regulated by surface markers associated with activation and senescence. CD40L, a costimulatory molecule, interacts with CD40 on B cells, promoting B cell proliferation and differentiation into antibody-producing cells and is commonly used as a marker of activation.^6,7,8,9^ Conversely, CD57 is associated with cellular senescence and terminal differentiation, a marker of limited proliferative potential and an increased susceptibility to apoptosis.^10,11^ These markers represent two ends of the Tfh cell functional spectrum, with CD40L indicating active, stimulatory capability and CD57 denoting cellular aging and senescence.

We previously found that the T cells of children in Bangladesh (an area of high microbial exposure) had increased effector and senescence-like phenotypes, which were not seen in age and sex matched children located in America (an area of low microbial exposure). This attribute was also associated with environmental factors such as stunting.^12^ However, the impact of specific environmental factors has yet to be studied in the context of T follicular helper cell function. This study investigates age-specific differences in Tfh cell CD40L and CD57 expression and subsequent humoral response post stimulation.

Environmental exposures, including infection, nutritional status, and breastfeeding practices, are known to shape immune development, particularly in early childhood.^13,14,15^ To understand how these factors influence Tfh cell functionality, we used Random Forest to identify important feature variables that influence cell responders. The covariates selected through Random Forest feature importance evaluation were used to fit a Generalized Estimating Equations (GEE) to assess the predictive power of specific environmental variables on CD40L and CD57 expression changes in Bangladeshi children. Furthermore, the functional state of Tfh cells has implications for immune outcomes, as these cells are integral to generating effective antibody responses post-vaccination or infection. To examine this outcome, we correlated CD40L and CD57 expression levels with antibody responses to routine vaccinations and Cryptosporidium infection, an endemic apicomplexan parasite in Bangladesh.

## RESULTS

### T follicular helper (Tfh) cells exhibit age-specific changes in functional marker expression in Bangladeshi children, at age 2

In both American and Bangladeshi children, the overall frequencies of CD4+ T cells and Tfh cells were comparable, suggesting that population-level differences in total T cell populations were minimal (**Fig. 1A**) and aligned with previous findings.^12^ However, significant differences in the expression patterns of CD40L and CD57 emerged in Bangladeshi children by age two, pointing to functional divergences in Tfh cell maturation or responsiveness. At age two, Bangladeshi children exhibited a diminished ability to upregulate CD40L following PMA/Ionomycin stimulation, a phenotype not observed in their American counterparts (**Fig. 1B**). This lack of CD40L upregulation signified a potentially reduced functional capacity of Tfh cells in supporting B cell responses. Additionally, Bangladeshi children at age two showed a significant decrease in CD57 expression upon stimulation, in contrast to American children, who did not exhibit this pattern (**Fig. 1C**), suggesting an accelerated apoptotic process in these cells. Collectively, these findings indicated that at age two, Bangladeshi children exhibited altered Tfh cell functionality, resembling an exhaustion-like phenotype potentially driven by environmental factors. This distinct immune profile was not observed in American children and may have implications for understanding immune competency across different environmental contexts.

**Figure 1.**
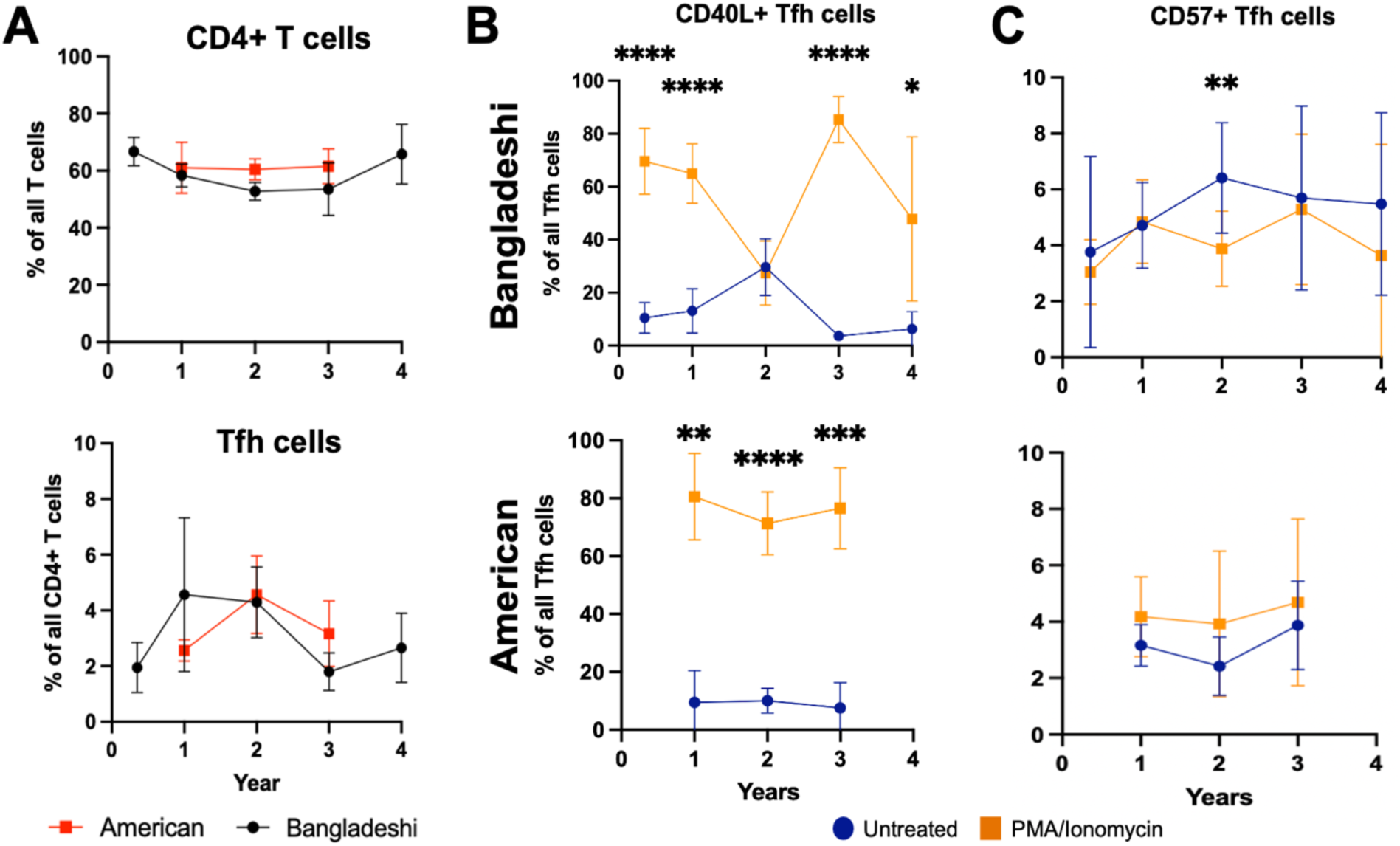
Comparison of CD4+ T cell and T follicular helper (Tfh) cell populations and function marker expression in American and Bangladeshi children. (A) Percentage of CD4+ T cells among all T cells and Tfh cells (CXCR5+, PD-1+) among all CD4+ T cells in American (red) and Bangladeshi (black) children across difference ages. (B) Percentage of CD40L+ Tfh cells out of all Tfh cells, comparing untreated (blue) and PMA/Ionomycin-stimulated (orange) conditions. (C) Percentage of CD57+ Tfh cells out of all Tfh cells, comparing untreated (blue) and PMA/Ionomycin-stimulated (orange) conditions. (B-C) Bangladeshi samples were displayed in the top panel and American samples in the bottom panel. Statistical significance was calculated by multiple paired T-test and denoted by asterisks (*p < 0.05, **p < 0.01, ***p < 0.001, ****p < 0.0001).

### Environmental factors predict expression changes of functional markers on T follicular helper cells upon stimulation, in Bangladeshi children at age 2

The variability in CD40L and CD57 expression on Tfh cells in Bangladeshi children, at age two, underscored the potential influence of environmental factors on immune cell functionality. Significant fluctuations in these markers upon stimulation revealed how external exposures may have uniquely shaped Tfh cell responses in this population (**Fig. 2**). To identify specific environmental factors associated with these functional changes, we evaluated feature importances with Random Forest and applied Generalized Estimating Equations (GEE) to assess the relative importance of external exposures and risk factors, including nutrition, infections, and breastfeeding practices influenced Tfh cell response to stimulation. Functional response to PMA/Ionomycin stimulation was measured by the ratio of percent CD40L+ or CD57+ Tfh cells post-stimulation to pre-stimulation levels.

**Figure 2:**
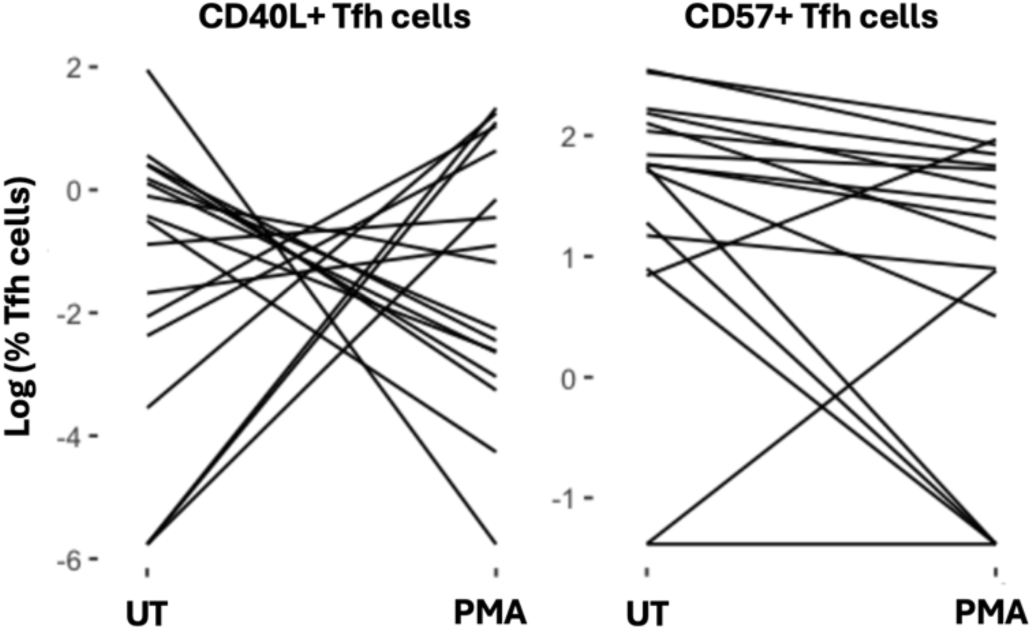
Variability in CD40L and CD57 expression on Tfh cells in response to stimulation in Bangladeshi children, at age two. Spaghetti plots show log-transformed percentages of CD40L+ (left) and CD57+ (right) Tfh cells before (UT) and after (PMA) stimulation with PMA/Ionomycin in individual Bangladeshi children, at age 2.

Key predictors identified included days of only exclusive breastfeeding, total number of antibiotic treatments by age 2, total number diarrheal episodes by age 2, and malnutrition scores such as weight-for-age Z-score (WAZ) (**Fig. 3A**). Partial dependency plots (**Fig. 3B**) further highlighted inverse relationships between these environmental factors and Tfh cell marker expression. Children who had longer exclusive breastfeeding, fewer antibiotic treatments, fewer diarrheal episodes, and better WAZ scores were more likely to upregulate CD40L expression upon stimulation. Conversely, children with more frequent antibiotic use, more diarrheal episodes, poorer malnutrition scores, and reduced exclusive breastfeeding duration were associated with increased CD57 expression. These findings suggested that early environmental exposures may influence Tfh cell functional programming, towards either an activation-supportive phenotype (CD40L+) or an exhaustion-prone phenotype (CD57+), with potential implications for immune competency and disease susceptibility in later life.

**Figure 3:**
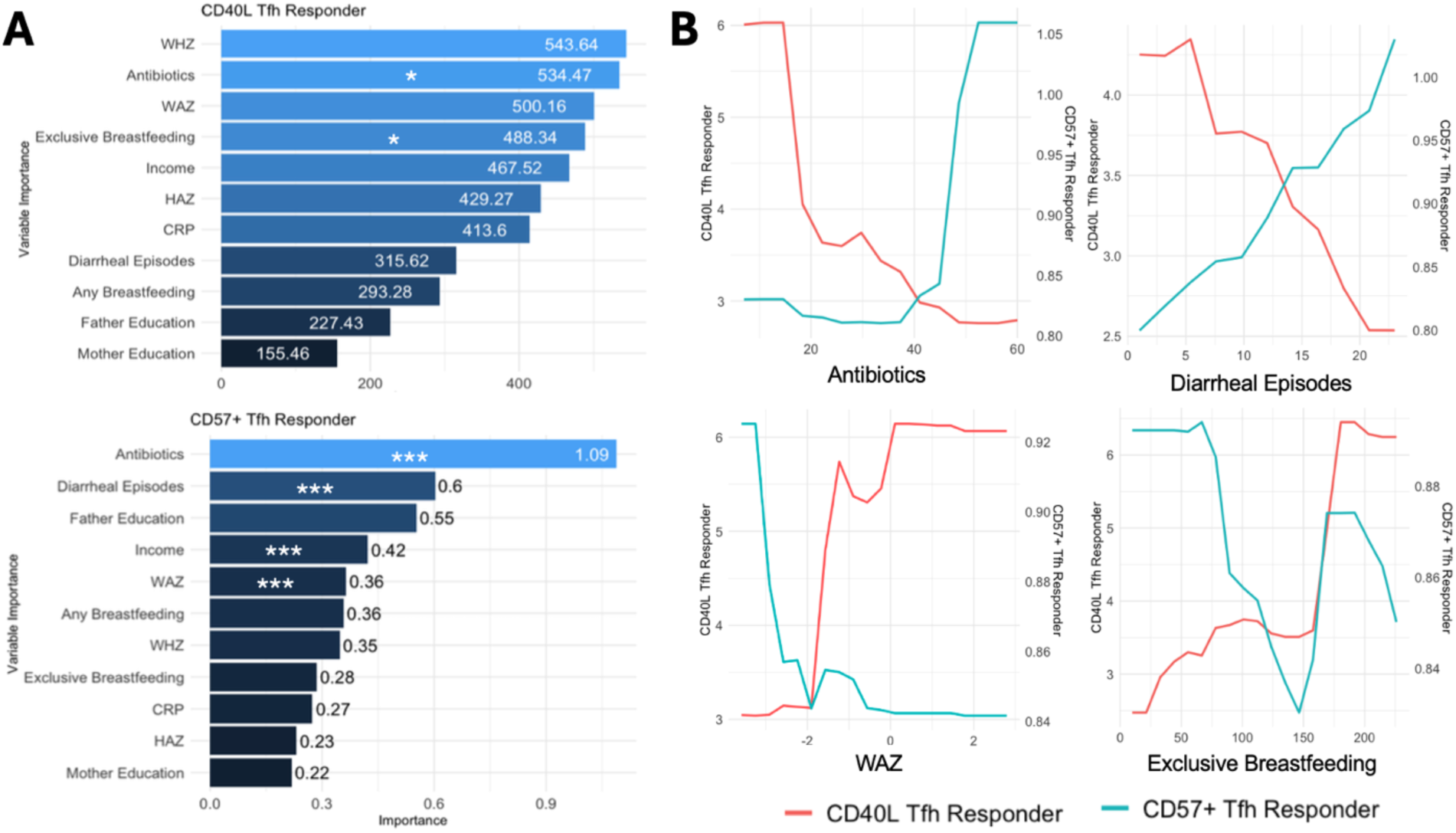
Environmental predictors of CD40L and CD57 expression in Tfh cells, at age 2, in Bangladeshi children. The response to PMA/Ionomycin stimulation was quantified by the percentage of CD40L+ or CD57+ Tfh cells after stimulation divided by their percentage before stimulation. Response to stimulation was then adjusted for a log scale distribution. (A) Variable importance plots generated by Random Forest model, ranking predictors of CD40L and CD57 response to stimulation (CD40L on the top left and CD57 on the bottom left). Variables are ranked by importance, with significant predictors determined by GEE model and marked by asterisks. (B) Partial dependency plots depict the relationship between key environmental factors (e.g., antibiotics, diarrheal episodes, WHZ, exclusive breastfeeding) and Tfh cell response, with CD40L response on the left y-axis (**pink**) and CD57 response on the right y-axis (**blue**). Statistical significance was calculated by GEE model and denoted by asterisks (*p < 0.05, **p < 0.01, ***p < 0.001, ****p < 0.0001).

### Functional T follicular helper cell responses are correlated with ability to mount an antibody response after vaccination or infection

To evaluate the relationship between Tfh cell functionality and downstream immune responses, we examined antibody concentration following vaccination or Cryptosporidium infection and correlated the increase of CD40L and CD57 expression on Tfh cells post-stimulation. Cryptosporidium, an endemic parasitic infection in Bangladesh, presented a relevant model for assessing pathogen-specific immune responses in this population.

Correlational analysis revealed that increased expression of CD40L and CD57 on Tfh cells in response to PMA/Ionomycin stimulation was associated with higher antibody titers against certain vaccine antigens, specifically Type 2 and Type 3 Poliovirus (**Fig. 4A**). These findings implied that the ability of Tfh cells to upregulate activation and senescence markers may have been most necessary to support robust antibody responses following Polio vaccination specifically. Additionally, we observed a significant positive correlation between high concentrations of IgA antibodies against the Cryptosporidium sporozoite protein Cp17 and CD40L expression. This indicated that Tfh cells expressing higher levels of CD40L may have played a role in promoting protective mucosal immunity against Cryptosporidium. Conversely, IgA levels against Cp17 were negatively correlated with CD57 expression, which suggested that Tfh cells with lower CD57 expression may have been more functionally active in mediating responses to parasitic infections (**Fig. 4B**).

**Figure 4:**
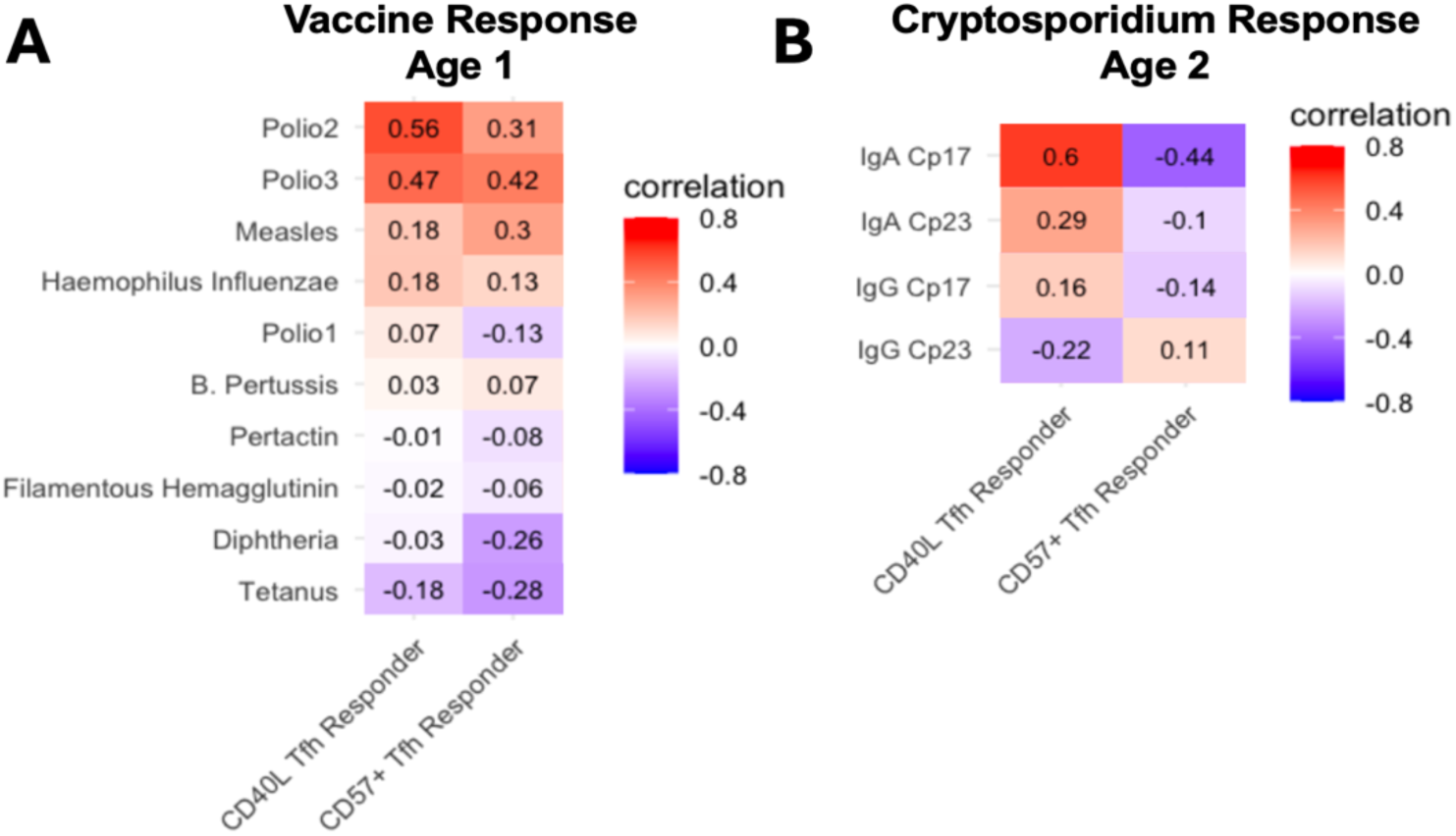
Correlation of Tfh cell functional response with antibody responses to vaccination and Cryptos-poridium infection. (A) Heatmap displaying correlation coefficients between Tfh cell CD40L and CD57 response to stimulation and antibody titers against vaccine antigens. (B) Heatmap displaying correlation of CD40L and CD57 response to stimulation with IgA and IgG responses against Cryptosporidium sporozoite proteins Cp17 and Cp23.

## DISCUSSION

This study demonstrates the significant impact of early-life environmental exposures on T follicular helper (Tfh) cell functionality in Bangladeshi children, with important implications for immune competency and humoral immunity. By focusing on CD40L and CD57 (markers of Tfh cell activation and senescence) we reveal age-specific and environment-driven alterations in Tfh cell responses that may compromise immune support for B cell-mediated antibody production.

At age two, some Bangladeshi children exhibit impaired CD40L upregulation and a significant reduction in CD57 expression upon stimulation. These findings suggest an exhaustion-like phenotype in Tfh cells that is not observed in American children of the same age. CD40L is critical for promoting B cell activation and germinal center formation, and impaired upregulation could hinder the generation of high-affinity antibodies and immunological memory. Meanwhile, the decreased CD57 expression upon stimulation may reflect accelerated apoptotic turnover of this population of Tfh cells, due to chronic immune activation.^16^ However, these findings vary between individual children, suggesting that environmental factors may be influencing the functional ability of Tfh cell response to stimulation.

Our analysis identified key environmental factors that predict Tfh cell functionality upon stimulation. Exclusive breastfeeding duration, number of antibiotic treatments, total diarrheal episodes, and malnutrition scores (WAZ) emerged as significant predictors of Tfh cell responses. Children with longer exclusive breastfeeding, fewer antibiotic treatments, higher WAZ scores, and fewer diarrheal episodes were more likely to upregulate CD40L, indicative of an activation-supportive phenotype. Conversely, frequent antibiotic use, poor nutritional outcomes, and recurrent diarrheal episodes were associated with increased CD57 expression, which signifies association between senescence or impaired Tfh cell functionality and chronic environmental stressors. These findings underscore the influence of early-life exposures on immune programming and highlight potential modifiable factors to improve immune health in high-risk populations. Interventions targeted to optimize immune development in resource-limited settings, such as promoting breastfeeding and improving sanitation to reduce diarrheal burden may promote Tfh cell function and potentially influence the ability to mount protective humoral responses post vaccination and infection.

In addition to examining environmental predictors, we investigated the downstream effects of Tfh cell functionality on humoral immunity. CD40L expression was positively correlated with IgA responses to the Cryptosporidium sporozoite antigen Cp17, indicating that Tfh cells with an activation-supportive phenotype may be important for effective immune responses. Conversely, CD57 expression was negatively correlated with anti-Cp17 IgA levels, supporting the hypothesis that a senescent or exhausted Tfh cell phenotype may be less effective in promoting humoral immunity. The positive association between CD40L expression and Cryptosporidium-specific IgA levels highlights the role of activation-supportive Tfh cells in mucosal immunity, particularly in environments with high pathogen exposure.

However, a different pattern emerged when examining the relationship between Tfh cell marker expression and antibody titers against vaccine antigens. Both CD40L and CD57 expression were positively correlated with increased antibody concentrations for Polio Type 2 and Type 3 antigens. This is most likely the result of the standard EPI vaccine administration in the first year of life. At age 1, Tfh cell response to stimulation was more homogenous in the Bangladeshi children, representing a healthier phenotype. It is possible that the results may have been more like that of the Cryptosporidium antibody response, in relation to CD40L and CD57 inverse correlation, if administered in the second year of life, when Tfh cells had an exhaustion-like phenotype. However, at age one, Tfh cell function may not yet have been fully shaped by environmental exposures to the extent observed at age two. As a result, increased CD57 expression upon stimulation at this earlier time point may not negatively impact the ability to mount a robust antibody response, contrasting with the patterns observed later in development.

Although limited in sample size, this study highlights the significant role of early immune programming in shaping long-term immune outcomes. The potential impact of an exhaustion-prone Tfh phenotype on susceptibility to infectious diseases and vaccine efficacy underscores the need for further research. These findings deepen our understanding of how environmental factors influence Tfh cell functionality and immunity and emphasize the importance of addressing environmental disparities to promote optimal immune health in children globally. Future research should focus on interventions to mitigate these environmental effects, such as targeted nutritional supplementation and improved access to healthcare, to enhance immune competence during critical windows of immune system development.

Additionally, the results emphasize Tfh cell functional markers, particularly CD40L and CD57, serve as valuable indicators of effective antibody-mediated immunity in response to vaccination and endemic infections. The distinct correlation patterns observed for CD40L and CD57 reinforce their roles in modulating the immune response and should continue to be studied, with CD40L upregulation associated with enhanced antibody production, while increased CD57 expression signaling a shift toward cellular senescence or diminished antibody support. These findings pave the way for leveraging Tfh cell markers to assess and potentially improve immune responses in populations at high risk for infectious diseases.

## METHODS

### Data Acquisition and Analysis

This study utilized data from two cohorts: PROVIDE of Bangladeshi children and STORK of U.S. children.

- PROVIDE Study: Based in Mirpur, Dhaka, Bangladesh, this cohort consisted of children from lower socioeconomic areas with high microbial exposure. Children were enrolled within seven days of birth and monitored biweekly. At each visit, questionnaires collected data on diarrheal illnesses, antibiotic use, breastfeeding practices (exclusive and non-exclusive), and nutritional status. Whole blood samples were collected annually up to age four. At study enrollment, mothers completed a lifestyle questionnaire. Vaccinations, including Polio (Types 1, 2, and 3), Haemophilus influenzae type B (Hib), Tetanus, Diphtheria, and acellular Pertussis, were administered before age one, while the measles vaccine was administered at 65 weeks of age.^17^
- STORK Study: Based in the San Francisco Bay Area, California, this cohort included children from areas with low microbial exposure. Pregnant mothers were recruited before 36 weeks of gestation and interviewed weekly post-birth. Whole blood samples were collected annually up to age three. Blood samples collected within 15 weeks of a child’s birthday were grouped together for analysis. For example, samples collected between 37–67 weeks of age were categorized as year 1.^18^

For all study samples, peripheral blood mononuclear cells (PBMCs) were isolated using conventional methods as previously described.^12^ Cryopreserved PBMCs from both studies were thawed, stimulated non-specifically with PMA/Ionomycin for six hours, and stained with metal-conjugated antibodies. Data acquisition was performed on a CyTOF2 mass cytometry instrument following established protocols. Detailed methodology for CyTOF immunophenotyping and analysis has been documented in detail,^12^ and the corresponding data files are publicly accessible through the Flow Repository database (Experiment ID: FR-FCM-ZYV8).

### Data Processing

CD4+ T cells were gated manually from the CD3+ T cell population following the exclusion of any dead cells, B cells, monocytes, and natural killer cells. Samples with fewer than a count of 400 CD4+ T cells were excluded from further analysis. Additionally, samples lacking matched untreated and stimulated groups were removed, ensuring that only paired untreated and stimulated samples were included in the final analysis. T follicular helper (Tfh) cells were gated from the CD4+ T cell population and defined as CXCR5+ PD-1+, CD4+ T cells. From the total Tfh cell population, CD40L+ and CD57+ Tfh cells were further gated out of the total Tfh cell population. The proportion of CD40L+ or CD57+ Tfh cells was calculated by dividing the count of CD40L+ or CD57+ Tfh cells by the total Tfh cell count within each sample, separately for untreated and stimulated groups. The Tfh cell response to stimulation was defined as the ratio of marker expression in the stimulated group to the untreated group. This ratio was calculated as the proportion of CD40L+ or CD57+ Tfh cells in the stimulated sample divided by the corresponding proportion in the untreated sample, providing a quantitative measure of Tfh functional response to stimulation.

### Prediction Modeling and Statistics

Random forest feature importance evaluation was used to identify key environmental factors influencing Tfh cell functionality. The variables of interest included environmental factors such as exclusive and total breastfeeding duration, number of antibiotic treatments, nutritional metrics (WAZ, HAZ, and WHZ scores), number of diarrheal episodes, CRP levels, annual family income, and parental education levels. The outcome variables were the log-transformed ratios of CD40L+ and CD57+ Tfh cells as a ratio of stimulated to unstimulated conditions. Models were trained using 10-fold cross-validation with tree counts of 100, 300, 500 and 700, and a minimum node sizes ranging from 1 to 9. Through the grid search, the optimal tuning parameters are selected to fit the Random Forest modeling. Variable importance was evaluated based on the mean decrease in the Gini index, which quantifies the predictive contribution of each variable.

Generalized Estimating Equation (GEE) models were subsequently employed to model associations between the top five predictors identified by Random Forest analysis. The log-transformed ratio of stimulated to unstimulated cell counts for CD40L+ and CD57+ Tfh cells (as a proportion of all Tfh cells) served as the response variables. GEEs accounted for the repeated measures and clustering inherent in the data, providing robust estimates of significant associations between environmental factors and Tfh cell functional outcomes. Statistical analyses were performed using appropriate software with significance thresholds set at *p* < 0.05.

### Antibody Quantification

Antibody concentrations post-vaccination for measles, *Bordetella pertussis* toxin, filamentous hemagglutinin, pertactin, diphtheria toxoid, tetanus, and *Haemophilus influenzae* type B antigens were quantified using standardized assays, reported as IU/mL, EU/mL, or ng/mL. IgA and IgG concentrations against Cryptosporidium sporozoite antigens Cp17 and Cp23 were measured using enzyme-linked immunosorbent assays (ELISA).

For ELISA, microtiter plates were coated with antigen solution and incubated overnight at 4°C. Plates were then washed, and a blocking buffer was applied to minimize nonspecific binding. Plasma samples were added to the wells and incubated, followed by another blocking buffer application. Subsequently, HRP-conjugated anti-human secondary antibodies were added to detect bound primary antibodies. TMB (3,3′,5,5′-tetramethylbenzidine) substrate solution was then added for color development, and the reaction was stopped with a stop solution. Absorbance was measured at 450 nm to quantify antibody levels. All measurements were performed in triplicate to ensure reproducibility.

**Table 1.** Samples used in the study.

|  | **Total**<br>**Sample #** | **18 weeks** | **52 weeks** | **104 weeks** | **156 weeks** | **208 weeks** |
| --- | --- | --- | --- | --- | --- | --- |
| **PROVIDE** | 75 | 14 | 26 | 23 | 6 | 6 |
| **STORK** | 22 | - | 5 | 9 | 8 | - |

## Data Availability

All data produced in the present study are available upon reasonable request to the authors. CyTOF data is available online in the Flow Repository database (Experiment ID: FR-FCM-ZYV8).

http://flowrepository.org/id/FR-FCM-ZYV8

## ACKNOWLEDGEMENTS

We thank Beth Kirkpatrick and Julie Parsonnet for their leadership roles in the PROVIDE and STORK studies respectively, and Uma Nayak for data management and the children and families for their participation. This work was supported by the Bill and Melinda Gates Foundation (OPP1113682) and NIH grant AI04356.

## BIBLIOGRAPHY

1. Deenick, E. K. et al. Follicular Helper T Cell Differentiation Requires Continuous Antigen Presentation that Is Independent of Unique B Cell Signaling. Immunity 33, 241–253 (2010).

2. Liu, R. et al. A regulatory effect of IL-21 on T follicular helper-like cell and B cell in rheumatoid arthritis. Arthritis Res. Ther. 14, R255 (2012).

3. Wang, J. et al. High frequencies of activated B cells and T follicular helper cells are correlated with disease activity in patients with new-onset rheumatoid arthritis. Clin. Exp. Immunol. 174, 212–220 (2013).

4. Chan, J.-A. et al. Age-dependent changes in circulating Tfh cells influence development of functional malaria antibodies in children. Nat. Commun. 13, 4159 (2022).

5. Oyong, Damian. A., et al. Adults with Plasmodium falciparum malaria have higher magnitude and quality of circulating T-follicular helper cells compared to children. EBioMedicine 75, 103784 (2021).

6. Essen, D. van, Kikutani, H. & Gray, D. CD40 ligand-transduced co-stimulation of T cells in the development of helper function. Nature 378, 620–623 (1995).

7. Whitmire, J. K., Slifka, M. K., Grewal, I. S., Flavell, R. A. & Ahmed, R. CD40 ligand-deficient mice generate a normal primary cytotoxic T-lymphocyte response but a defective humoral response to a viral infection. J. Virol. 70, 8375–8381 (1996).

8. Conley, M. E. et al. Primary B Cell Immunodeficiencies: Comparisons and Contrasts. Annu. Rev. Immunol. 27, 199–227 (2009).

9. Noelle, R. J. et al. A 39-kDa protein on activated helper T cells binds CD40 and transduces the signal for cognate activation of B cells. Proc. Natl. Acad. Sci. 89, 6550–6554 (1992).

10. Brenchley, J. M. et al. Expression of CD57 defines replicative senescence and antigen-induced apoptotic death of CD8+ T cells. Blood 101, 2711–2720 (2003).

11. Elias Junior, E., et al. CD57 T cells associated with immunosenescence in adults living with HIV or AIDS. Immunology 171, 146–153 (2024).

12. Wagar, L. E. et al. Increased T Cell Differentiation and Cytolytic Function in Bangladeshi Compared to American Children. Front. Immunol. 10, (2019).

13. Chandra, R. Nutrition and the immune system: an introduction. Am. J. Clin. Nutr. 66, 460S–463S (1997).

14. Oddy, W. H. The impact of breastmilk on infant and child health. Breastfeed. Rev. Prof. Publ. Nurs. Mothers Assoc. Aust. 10, 5–18 (2002).

15. Kang, T. G. et al. Epigenetic regulators of clonal hematopoiesis control CD8 T cell stemness during immunotherapy. Science 386, eadl4492 (2024).

16. Palmer, B. E., Blyveis, N., Fontenot, A. P. & Wilson, C. C. Functional and Phenotypic Characterization of CD57+CD4+ T Cells and Their Association with HIV-1-Induced T Cell Dysfunction1. J. Immunol. 175, 8415–8423 (2005).

17. Kirkpatrick, B. D. et al. The “Performance of Rotavirus and Oral Polio Vaccines in Developing Countries” (PROVIDE) Study: Description of Methods of an Interventional Study Designed to Explore Complex Biologic Problems. (2015) doi:10.4269/ajtmh.14-0518.

18. Ley, C. et al. Stanford’s Outcomes Research in Kids (STORK): a prospective study of healthy pregnant women and their babies in Northern California. BMJ Open 6, e010810 (2016).

